# Implementation of an internet-based stress management program in micro- and small-sized enterprises: A study protocol for a pre-post feasibility study of the effectiveness-implementation hybrid type 2 trial

**DOI:** 10.1101/2023.11.02.23297716

**Authors:** Natsu Sasaki, Taichi Shimazu, Hajime Takeno, Sayaka Ogawa, Utako Sawada, Akizumi Tsutsumi, Kotaro Imamura

## Abstract

**Background:** Text-based self-guided internet-based stress management programs can improve mental health among workers. However, such mental health programs have scarcely been implemented in micro– and small-sized enterprises (MSEs), which are lacking in occupational healthcare services.

**Objectives:** This paper describes a study protocol for a pre-post feasibility study of an effectiveness-implementation hybrid type 2 trial of text-based internet-based programs (“WellBe-LINE”) in MSEs with less than 50 employees. This feasibility study primarily aims to evaluate trial methods for future effectiveness-implementation hybrid type 2 trials.

**Methods:** For this study protocol, an internet– and text-based self-care intervention program using the LINE app (a popular message tool in Japan) will be prepared according to evidence-based psychoeducational topics. Based on our online survey findings, personalized algorithms will be implemented according to employees’ gender, age, and psychological distress levels. A pre-post design feasibility study will be conducted on ten companies to evaluate trial methods (e.g., recruitment, penetration, and procedures). The primary outcome will involve individual-level penetration, defined as the proportion of the number of employees who register for the program divided by the total number of invited employees at the company. The progression criterion specifies that more than 50% of the recruited companies obtain 60% individual penetration. Finally, acceptability, appropriateness, feasibility, and cost will be measured using quantitative internet-based questionnaires and qualitative interviews.

**Discussion:** This pre-post feasibility study for future effectiveness-implementation hybrid type 2 trials will provide in-depth knowledge about the successful implementation of text-based, semi-personalized, self-care mental health interventions in real-world settings using both quantitative and qualitative data.

**Conclusions:** This feasibility study will help validate the effectiveness of text-based interventions using a widely used social networking service (SNS) tool for employees in MSEs.

**Trial registration:** UMIN clinical trial registration: UMIN000046960 (registration date: February 21, 2022)

https://center6.umin.ac.jp/cgi-open-bin/ctr/ctr_view.cgi?recptno=R000053570

**Contributions to the literature:**

- Internet-based mental health interventions in micro– and small-sized enterprises have not been implemented yet.
- This feasibility study plans to test the implementation strategies to achieve high penetration in employees.
- This study can provide insights into occupational health implementation in a disadvantaged context.

## Background

Worldwide, mental health problems in the workplace impact individual health-related disability and productivity loss (1, 2). Micro-, small-, or medium-sized enterprises (MSMEs) are important targets for mental health interventions (3) as they represent about 90% of businesses and more than 50% of employment worldwide (4). However, MSMEs, especially micro– and small-sized enterprises (MSEs) with fewer than 50 employees, are less likely to implement health promotion programs because of limited resources, such as cost, access, and time (5). Thus, offering evidence-based programs alone may not be valuable for employees. While implementing the primary prevention of mental health in MSMEs is challenging (3), preventive interventions for mental health is generally more effective when used by a large populations (6). Therefore, increasing the adoption and penetration (reach) of health promotion programs are key objectives. To achieve high penetration, interventions and strategies specific for MSEs must consider implementation barriers.

Previous studies have identified barriers to the implementation of mental and health interventions in small companies (7–12): low leadership engagement of employers (7), lack of knowledge about the impact on business (8), limited resources (9, 12), limited time and money (11), regulations (no legal requirement) (12), lack of understanding on the necessity of the interventions and suboptimal approach (10). Additionally, job-related stressors between small and large companies varied (13). In small companies, poor psychosocial factors at work and poor communication are strongly associated with job stress (13). Therefore, personalized intervention content and specific delivery strategies to overcome organizational barriers in MSEs are needed.

Internet-based interventions may solve problems of implementing mental health interventions in MSEs. Internet-based intervention is feasible, cost-effective, and accessible (14–16), meeting the needs of MSEs. A comparison of face-to-face and internet-based interventions revealed no differences in their effectiveness in treating common mental disorders (17, 18). Despite the small effect size, even text message-delivered interventions have been proven to be effective for stress management (19).

Moreover, tailoring (or personalizing) messages in internet-based interventions proving even more effective in stimulating changes in health behavior (20). Internet-based and personalized text message interventions may thus be suitable for improving the acceptance of employees, leading to successful implementation in MSEs. Acceptability, appropriateness, and feasibility have been suggested as important implementation aspects of mental health interventions (21–24). However, few studies have not been conducted yet examined the effectiveness and implementation outcomes of internet-based interventions for MSEs without some recent protocols (25, 26). Therefore, methods to improve implementation outcomes (i.e., implementation strategy) should be developed and examined.

As MSME managers have little motivation to implement the program (5, 7), a specific implementation strategy personalized to the context should be adopted. According to the Expert Recommendations for Implementing Change (ERIC), educating stakeholders (employees, employers/managers, and recruiters) is a possible strategy if leadership engagement can be a barrier in the context (27, 28).

Informing employers about the importance of preventive measures in mental health and effective procedures for introducing the program to employees may increase their adoption. However, no study has investigated the effect of internet-based interventions on MSEs with such implementation strategies (i.e., educating employers). Previous implementation research for mental health interventions using the internet or technologies suggested the negative attitude of stakeholders (and, particularly, the users) for internet-based interventions (more preference for face-to-face) (29, 30) and require education for providers (31)

Hybrid-type designed studies can be conducted between effectiveness studies and implementation research (32). The hybrid type 2 design can be used to test the effectiveness in general practice settings without controlling/ensuring delivery of the intervention and implementation process (32). Moreover, feasibility and pilot studies (33) can help build and test effective implementation strategies by addressing uncertainties around design and methods and identifying potential causal mechanisms (34). According to the CONSORT 2010 statement: Extension to randomized pilot and feasibility trials, pilot or feasibility studies should have clear criteria for deciding whether to progress to the next full trial (35). Because there are still many uncertainties in MSEs settings and less evidence is available in implementation research, a pilot feasibility study is important to obtain insights about how we should act in the next stage.

### Aims and objectives

This pre-post feasibility study on a future effectiveness-implementation hybrid type 2 trial aims to evaluate the implementation trial procedures of text-based online programs in micro– and small-sized enterprises with less than 50 employees.

The objectives of this feasibility study, in preparation for a future trial, are as follows:

1. To evaluate the feasibility of trial methods and strategies by setting individual-level penetration as an indicator of successful implementation (primary outcome).
2. To evaluate the feasibility of recruitment, procedures, and strategies for disseminating the program.
3. To evaluate the acceptability, appropriateness, fidelity, and cost, among recruiters, employers/managers, and users.
4. To ensure no harm on psychological distress of the users.

## Methods/Design

### Program outline of “WellBe-LINE”

The text-based self-care intervention program “WellBe-LINE,” which was customed for employees in MSEs, will be set in the LINE app (commonly used SNS chat tool in Asian countries) based on evidence-based psychological contents. One text message will be sent once per week and includes a website link (URL) for more information. **Figure 1** presents an outlook on the message. The portal website (https://wellbeing-kokoro.com/) contains more than 100 articles on mental health, comprising topics about problem solving (36), acceptance and commitment therapy (37), self-compassion (38), sleep hygiene (39), cognitive behavioral therapy for insomnia (40), and physical activity (41). **Figure 2** presents the procedures for starting the program and the intervention schedules.

**Figure 1.**
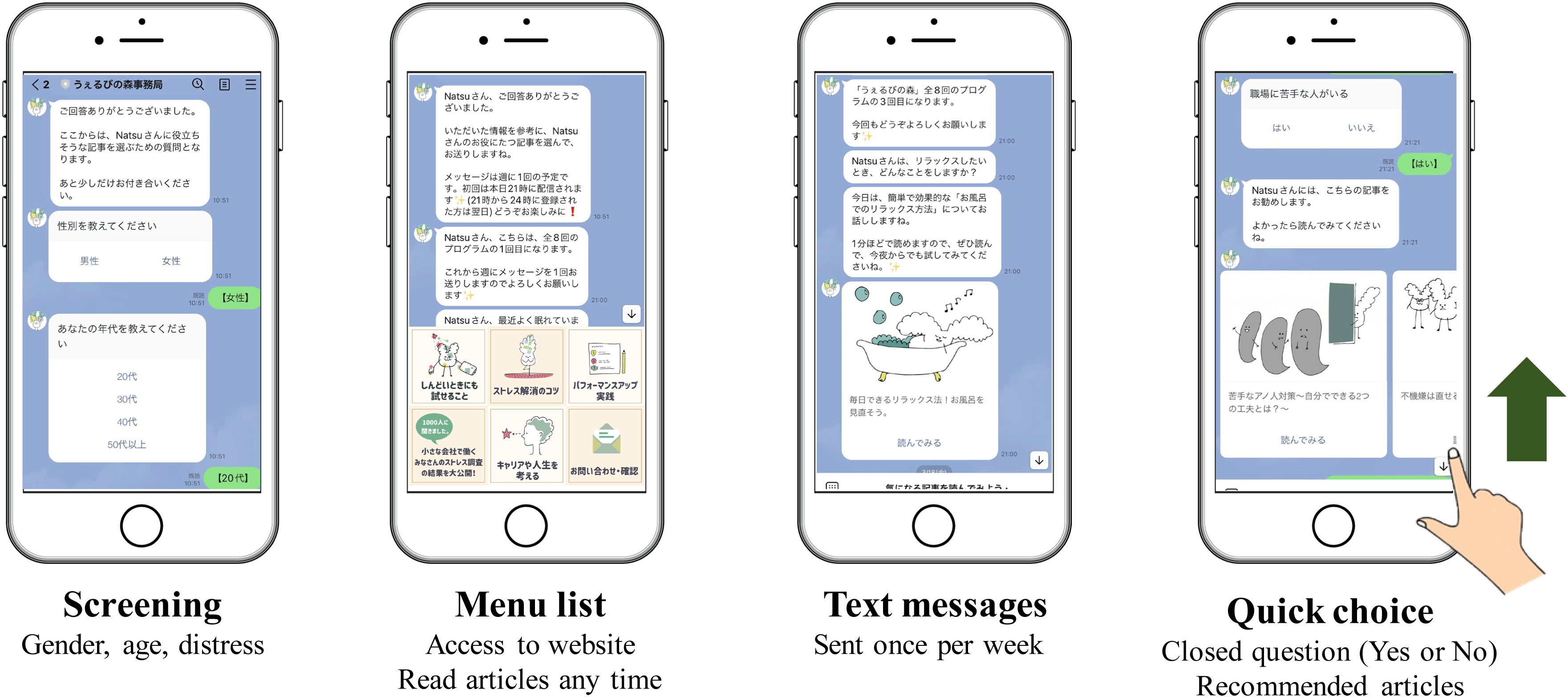
Outlook of “WellBe-LINE”.

**Figure 2.**
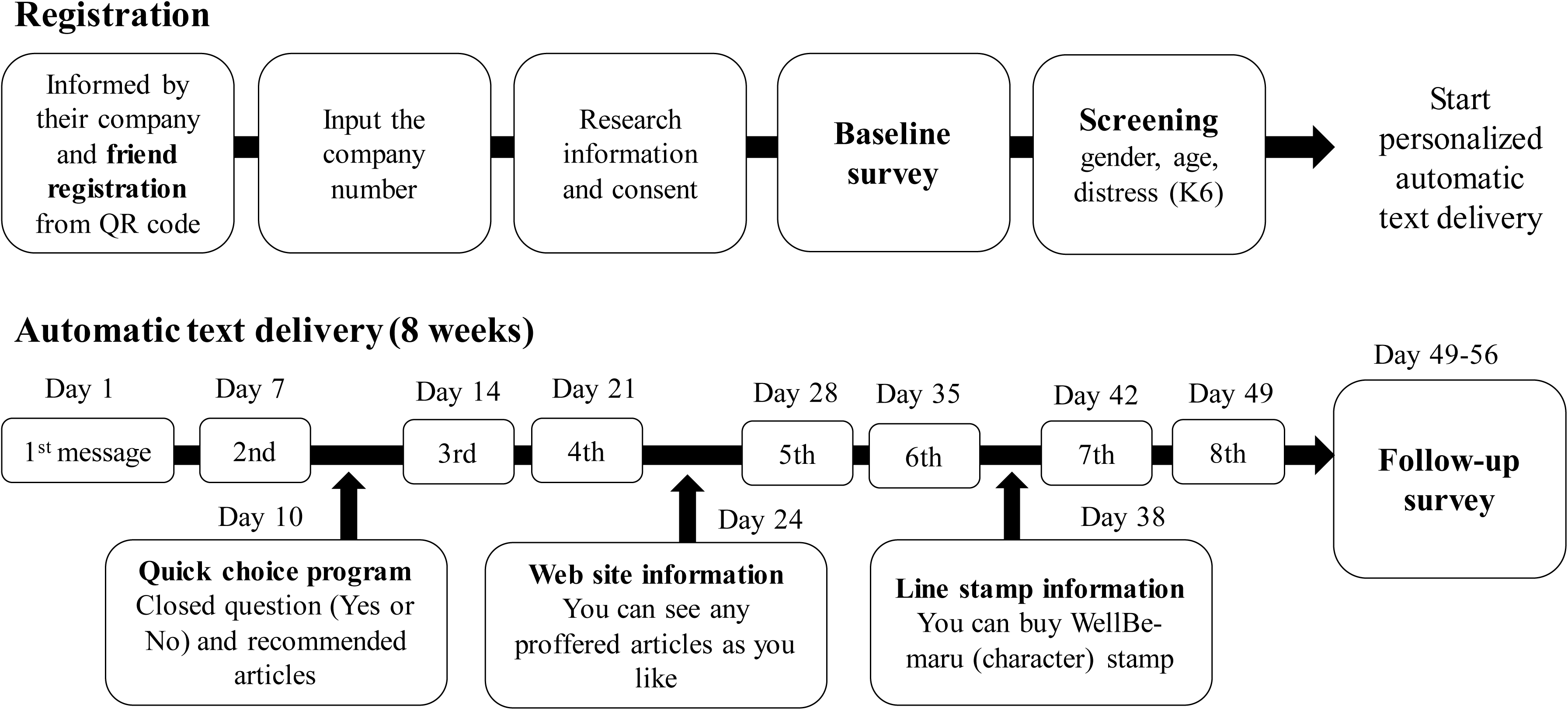
Procedures of starting “WellBe-LINE” and schedule of the intervention.

### Personalization

Personalizing programs are effective at creating changes in health behavior (20), maximizing effectiveness despite the low intensity of the program. We provide 16 scenarios according to the 16 groups (gender*four age category*psychological distress [K6 ≥ 5, or K6<5]) by screening check when participants started the program. **Supplementary File 1** presents Detailed information about the personalized program.

“WellBe-LINE” provides messages in order of rank, from highest to low (week 1 to 8). **Supplementary File 2** reports the final personalized scenario for the 16 groups.

### Study design

In the feasibility study, A one-repeated-measure (pre-post) design will be used. Measurements will be taken at baseline (pre) and 8 weeks afterwards (post). Data collected will then be used to further refine the intervention, recruitment, procedures, strategy for disseminating, and power of a subsequent cluster RCT, which is a hybrid type 2 trial. This study protocol was approved by the Research Ethics Committee of the Graduate School of Medicine, Faculty of Medicine, University of Tokyo (2021190NI-(1)). The trial registration is available elsewhere (UMIN000046960).

### Participants

At the organizational level, companies with fewer than 50 employees will be recruited. At the individual level, employees over 18 years old will be recruited, regardless of their employment contract (e.g., part time). As this study examines the implementation of preventive programs for all employees, no further exclusion criteria is needed.

### Recruitment and procedure

To activate MSEs’ employers/managers, we will use a pre-existing trustful pipe with the MSEs. Five licensed social insurance consultants (recruiters), who were Patient Public Involvement (PPI) members, recruited MSEs that they had a relationship with. recruiters provide the research information and invitation to demo program (short version of “WellBe-LINE” which finish one week). If the invited company is interested in participating in the research, the recruiter will provide contact information for researchers. The researchers will then coordinate with the employers/managers on the date of the online meeting (described in the next paragraph).

After the meeting, researchers provide one poster and one original clear file for each employee. Using a template, researchers will also e-mail employees to invite employees to participate in the program. The researcher will provide one “code” for each company. Employers/managers inform employees about the present research by the way they fit the company (e.g., e-mail, posters, and other ways of appealing them). Employees who participate in the program must enter the code after registering for the program via their personal LINE account. Research details will then be explained on the LINE chat, and they will provide their concert by pushing the button to start answering the baseline survey. Employees select their sex and age categories. They also answer six questions about psychological distress (measured by K6). The program automatically recognizes their characteristics (sex × age × distress [5+ high or low]) and starts the program.

### Online meeting with employers/managers (implementation strategy)

Licensed social insurance recruiters engage in the recruitment process to motivate relevant employers and managers to participate in the study. After obtaining agreement, researchers will provide 30–45 minutes of online semi-structured lectures (meetings) with employers and managers via zoom to determine the situation at the company and provide knowledge about behavioral changes and an effective way to invite employees for participation and attain high penetration. A previous qualitative study suggested that fundamental factors influencing the implementation of workplace health promotion in SMSEs entail the leadership engagement of employers (7). **Table 1** presents and justifies the contents of the semi-structured lecture with implementation strategy specification guidelines introduced by Proctor et al. (42) for each component. Semi-structured lectures will then be conducted using Microsoft PowerPoint. Psychological techniques for engaging employees are based on classical behavioral psychology related to motivation theory (43–46) and processing fluency related to health information (47).

### Measurements

Outcomes will be measured using online self-report questionnaires and interviews with participants (employees), employers/managers, and licensed social insurance consultants (recruiters). **Table 2** presents outcomes measured for each stakeholder.

#### Implementation outcome

##### Penetration (primary)

Individual-level penetration is calculated as the proportion of the number of employees who register for the program divided by the total number of employees at the company. The progression criterion (35) is that over 50% of the recruited companies obtain 60% individual-level adoption. The rationale for setting this goal is that from the results of an online survey of 1000 employees at MSEs, we asked the question, “If the company you work for provided you with information about the program registering as a friend on LINE regarding information useful for mental health and work, would you be willing to do so?” Specifically, 44.8% of the respondents answered “Yes or “Fairly agree” to the question. For the second question, “Please select one app or social networking site that you are most likely to use (or most prefer) when receiving mental health or work-related information.” Meanwhile, 35% of the respondents answered “do not want to receive such self-care information.” From these results, it is assumed that the estimated participation ratio would be 45-65%, and that 60% of the company’s own employees would be registered as friends on LINE, which would be considered a high level of penetration at the individual level. If we do not achieve the progression criteria [32], the entire process and delivery of the program will be revised based on the findings of the present feasibility study.

##### Acceptability, appropriateness, and feasibility

Acceptability, appropriateness, and feasibility were measured using questionnaires and interviews with employees (users) and employers/managers. Implementation outcome scales for digital mental health (iOSDMH) will be used in a questionnaire to assess three domains of implementation outcomes (24). The iOSDMH has three versions (i.e., users, providers, managers, and policymakers). The items of the iOSMDH were developed through a literature review, and the outcome was organized according to Proctor’s implementation outcomes (22). The response options include a four-point rating for users and five-point rating for providers, and for managers or policy makers, with the added option of “Don’t know.” In this study protocol, employees will then be asked to use iOSDMH for users and employers/managers by iOSDMH for providers. If employers and managers (or people who are concerned and provide the program in the company) are clearly separated, iOSDMH will be used by managers, policymakers, and providers, respectively.

##### Fidelity

Fidelity will be assessed through interviews with employers/managers and licensed social insurance consultants (recruiters) about the implementation strategy. The authors will provide them with materials for the invitation of employees and companies. Although the method of recruiting can be personalized according to the user’s context and local culture, in post interviews (T2), the extent to which they made arrangements deviated from the originally planned way of invitation will be assessed.

##### Cost

Cost will be assessed through interviews with employers/managers and licensed social insurance consultants (recruiters). The time and money spent on program implementation will be assessed using a questionnaire. Additionally, whether the reward from the implementation matches cost will be assessed. Participating organizations will not incur direct implementation costs in “WellBe-LINE” and will not be charged for the use of the program during the study.

#### Health outcome

##### Users’ psychological distress

Psychological distress was measured using K6 (Kessler 6) (48, 49). Respondents will be asked to report how frequently they had experienced the following six symptoms in the past 4 weeks: felt nervous, hopeless, restless or fidgety, worthless, depressed, and felt that everything was an effort. Response options included “none of the time,” “a little of the time,” “some of the time,” “most of the time,” and “all of the time. Scores range from 0 to 24. The Japanese version of the K6 exhibits good reliability and validity (50). K6 works well as the CIDI Short Form in identifying cases of clinically significant mental disorders (51). Scores over 5 are judged as having moderate psychological distress and can be used as a cutoff point for high or low distress (52, 53).

##### Work performance (HPQ)

Work performance will be evaluated using one item of the WHO Health and Work Performance Questionnaire (HPQ) (54). Participants will then be asked to rate their overall work performance over the past four weeks. Items were scored on an 10-point scale ranging from 0 (worst) to 10 (best), with high scores indicating good work performance. The Japanese version of the HPQ exhibits good reliability (55).

##### Work engagement (UWES-3)

The ultra-short form of the Utrecht Work Engagement Scale, three items (UWES-3) will be used to assess work engagement (56). The UWES-3 consists of three subscales (i.e., vigor, dedication and absorption) with each of one item. The UWES-3 is a self-reported seven-point rating scale (0 = never; 6 = everyday). The mean score of the three UWES subscales, and total score is computed by adding the scores and dividing the sum by the number of items in each subscale. The Japanese version of the UWES-3 exhibits good reliability and validity (57).

##### Job satisfaction

Job satisfaction will be measured using one item from the Brief Job Stress Questionnaire (BJSQ) (58) on a four-point Likert scale. Higher scores indicated higher job satisfaction.

##### Euthymia

Euthymia is a transdiagnostic construct for well-being and represents psychological flexibility, a unifying outlook on life, and resistance to stress (59, 60). The euthymia Scale (ES) is a 10-item index with dichotomous options (False = 0; True = 1). This results in total scores ranging from 0 to 10, indicating better euthymic state for higher scores. The Japanese version of the ES has high concurrent validity and sensitivity as a clinimetric scale (61).

##### Good relationship with co-workers

The original one-item measurement (“I can work well with my colleagues”) will be used to assess relationships with co-workers on a four-point Likert scale.

##### Trust management

Trust in management is measured by one item (“I can trust with the management of my organization”) from the new BJSQ (62) on a four-point Likert scale.

##### Cares about employees

The following original one-item measurement will be included at the request of the PPI partner: “Using company research funds, the Wellbe-LINE program was developed to support the mental health of people working at companies with less than 50 employees. How much did the introduction of this program make you feel that your company cares for its employees?” A four-point Likert scale is used for measurement.

#### Process outcome

##### Block rate

LINE accounts can reject or “block” contact from other accounts. We will assess this rate of blocking by system detection.

##### Uncomfortable experience/harms

Uncomfortable experience/harm will be assessed using five items from the iOSDMH (24). Time consumption, mental symptoms, induced dangerous experiences regarding safety, physical symptoms, and excessive pressure on regular learning will be measured for employees (users) in the questionnaire. Employers/managers and recruiters will be asked to rate the following item: “This program does not result in negative side effects (e.g., physical or psychological symptoms).” The response options were four-point rating scales for users and five-point rating scales for providers (managers or policy makers), with the added option of “Don’t know.”

##### Adherence (completion rate)

Whether employees read or engaged with messages could not be accurately detected in LINE owing to technical limitations. Instead, we asked employees to answer four questions related to adherence in the follow-up survey: “How many times did you read the LINE message?,” “How many times did you visit the website?,” “Did you visit other websites when you received a notification on LINE?,” and “Did you read other related articles in the website?.”

#### Analysis

##### Sample size

A previous review suggested that 25 employees are needed if the effect size is likely to be moderate to large to test the efficacy (63). However, this study protocol aims to determine sample size. We set the target sample size to 10 companies (200 employees) based not on the accuracy of the results (e.g., 95% CI) but on the feasibility of relevant resources. This study protocol will support our decision to proceed to the next large trial and will be utilized for our revisions to the program and implementation strategy.

#### Quantitative data

Health outcomes will be evaluated using a paired t-test between pre-(T1) and post-intervention (T2) according to protocol. The effect sizes and 95% CIs were calculated using Cohen’s d only for those who completed the post-intervention questionnaire. Statistical significance was defined as P < 0.05. IBM SPSS Statistics® version 28 was used for all analyses.

#### Analysis of qualitative data

To adapt the implementation strategy (i.e., online meetings with employers/managers) and program into the real context, descriptive data from the interview will be summarized according to the Consolidated Framework for Implementation Research (CFIR) (64). We will describe the process and experience of making decisions for future cluster RCT.

#### Patient and Public Involvement (PPI)

PPI facilitates enhanced quality, appropriateness of research, and development of user-friendly research materials (65). In our study, seven PPI partners knew about the real context of MSEs: four licensed social insurance consultants, one occupational health physician who provided services in MSEs, one employer in MSEs (one of co-authors, HT), and one manager in the Tokyo Chamber of Commerce and Industry who organized the “Health and Productivity Management.” These PPI partners have participated in all research stages, including the program development, user-relevant research questions, user-friendly materials, more appropriate recruitment strategies for studies, discussion of the interpretation of data, and dissemination of study results. The PPI process is described based on the PPI handbook and reporting checklists (66, 67).

## Discussion

This study protocol paper aims to evaluate implementation outcomes via the “WellBe-LINE” in MSEs with less than 50 employees. “WellBe-LINE” is a text-based, semi-personalized, self-care mental health intervention program through the LINE app. The program sends messages with content considered effective at preventing mental issues (36–41). This pre-post feasibility study for future effectiveness-implementation hybrid type 2 trials will provide in-depth knowledge about the successful implementation of self-care interventions in real-world settings using both quantitative and qualitative data. Any adverse events and/or unsuccessful procedures will be utilized to identify alternative strategies before proceeding to the next full trial.

This study protocol will apply the specific implementation strategy wherein researchers meet with employers/managers online for 30–45 minutes to encourage them to invite employees to participate in the “WellBe-LINE” Program. This strategy is based on previous findings of leadership engagement being a considerable barrier in the health program implementation (7). Furthermore, PPI partners who engaged in all research stages will provide essential suggestions in implementation, leading to a positive impact on field-specific studies (65).

### Limitation

This study protocol has several limitations. First, the sample size may not be adequate to detect significant effectiveness in reducing psychological distress owing to the pilot nature of this study. Second, this study protocol lacks a control group; the true impact of the implementation strategy can only be determined in the next step. Third, generalizability is limited owing to the recruitment process. Hence, MSEs highly interested in health promotion should be included in this study. However, this study protocol primarily aims to test the implementation outcomes, including adoption and feasibility, in the accessible sample. Future studies could engage and recruit MSEs with high heterogeneity may be required in the. Fourth, individual-level adoption may be affected by personal beliefs about mental health issues in the workplace (e.g., social desirability(68), fear of stigmatizations(69), concerns about negative impact at the workplace(70)). Thus, factors not related to the implementation strategy can be considered if the outcomes are evaluated.

## Conclusion

MSEs are less likely to implement health promotion programs worldwide, although many employees work there. The low-cost and easy-to-access self-care program “WellBe-LINE” targeted specifically at the needs of MSEs can be a solution, despite its low intensity, if the penetration of employees is applicable. The wide range of evaluations proposed in this study protocol will provide valuable suggestions for implementing preventive health promotion measures in MSEs.

## Declarations

### Ethics approval and consent to participate

This study protocol was approved by the Research Ethics Committee of the Graduate School of Medicine/Faculty of Medicine, The University of Tokyo, No.2021190NI-(1).

## Consent for publication

Not applicable.

## Availability of data and materials

The data related to the present study protocol are available from the corresponding author, KI, upon reasonable request.

## Authors’ contributions

KI was in charge of this study protocol, supervised the process, and provided his expert opinion. AT provided expert suggestions regarding the study concept as a supervisor. NS and KI organized the study design. SO and US supported the study management. HT analyzed the pilot data and created an algorithm for the program. Collaborator TS provided his expert opinion from the viewpoint of implementation science and ensured that questions related to the accuracy or integrity of any part of the work were appropriately investigated and resolved. NS and KI wrote the first draft of the manuscript, and all the other authors critically revised the manuscript. All the authors approved the final version of the manuscript.

## Abbreviations

BJSQ: Brief Job Stress Questionnaire
MSEs: Micro-, or small-sized enterprises
MSMEs: Micro-, small-, or medium-sized enterprises
PPI: Patient and Public Involvement

## Acknowledgements

This study protocol was supported by the National Center Consortium in Implementation Science for Health Equity (N-EQUITY) funded by the Japan Health Research Promotion Bureau (JH) Research Fund (2019-(1)-4) and JH Project Fund (JHP2022-J-02).

