## Supplemental table for "Implementation of an internet-based stress management program in micro- and small-sized enterprises: A study protocol for a pre-post feasibility study of the effectiveness-implementation hybrid type 2 trial"

**Supplementary file 1. Detailed information on how to personalize the program (“WellBe-LINE”).**

To determine user interest in mental health and job-related topics, we conducted an online survey of 1000 employees in MSEs (recruited equally by gender and age). We asked three questions: “What do you want to know about workers’ mental health (up to 5)?” with 14 response options (e.g., stress reduction and relaxation), including “Nothing I want to know”; “What do you want to know about workers’ situation other than their mental health (up to three options)?” with eight response options (e.g., career, work-life balance), including “Nothing I want to know”; and “What do you want to know about work (up to three options)?” with eight response options (performance improvement, communication), including “Nothing I want to know.” Levels of interest in the items in the questionnaires were scored and ranked for delivering articles with higher interest in each of the 16 groups (gender × four age categories × psychological distress [K6 ≥ 5, or K6<5]). Moreover, the number of selected responses for each item per selected item per group was used as a score (n). Because the selectable item number in the questionnaire was different among the three questionnaires, the possible selection of one item was different. Therefore, the scores were normalized by the ratio of the selectable item number to the total number of items in the questionnaire (R). In addition, there was no correlation among the three questionnaires to be compared, and the scores were calibrated by using one item “Nothing I want to know,” which was common in all questionnaires. We utilized the reciprocal of the score in the item to show how high the respondent’s interest was in a group and put the reciprocal as a standard to be compared (C).

. The calibrated scores (S) are given as

$S_{ij}=\frac{N_{ij} * C_{i}}{R_{i}}=$ $\frac{n_{ij} * \mathrm{Tot}_{i}}{{{Nt}_{i}* Sel}_{i}}.$

S = Calibrated scores as the levels of interest

i = Id of a questionnaire

j = Id of an item in the questionnaire

N = n / Ts

n = Number in a selected item

Ts = Total of selected items in a questionnaire in a group

R = Sel / Tot

Sel = Number of items that one respondent can select in a questionnaire

Tot = Number of total items in a questionnaire

C = value for calibration = Nt / Ts

Nt = Number of selected responses in the item of “nothing interesting in the questionnaire”

**Supplementary file 2. Personalized scenario for 16 groups by gender, age (four categories), and psychological distress.**

|  | Low distress (K6<5) | | | | | | | |
| --- | --- | --- | --- | --- | --- | --- | --- | --- |
|  | Men | | | | Women | | | |
| Rank | Age 20–29 | 30–39 | 40–49 | Over 50 | Age 20–29 | 30–39 | 40–49 | Over 50 |
| 1 | Job performance | Better sleep | Better sleep | Better sleep | Better sleep | Better sleep | Better sleep | Better sleep |
| 2 | Work environment | Stress reduction | Job performance | Relaxation | Relaxation | Relaxation | Relaxation | Relaxation |
| 3 | Career | Relaxation | Good lifestyle | Stress reduction | Motivation | Stress reduction | Good lifestyle | Job performance |
| 4 | Stress reduction | Motivation | Stress reduction | Good lifestyle | Positive feeling | Good lifestyle | Stress reduction | Good lifestyle |
| 5 | Better sleep | Job performance | Work environment | Job performance | Job performance | Work-life balance | Stress coping | Mindfulness |
| 6 | Happiness | Positive feeling | Work-life balance | Work-life balance | Stress reduction | Work stress | Mindfulness | Positive feeling |
| 7 | Work-life balance | Career | Work engagement | Work environment | Good lifestyle | Job performance | Problem solving | Stress reduction |
| 8 | Work engagement | Work environment | Motivation | Positive feeling | Work-life balance | Positive feeling | Job performance | Communication |
|  | High distress (K6≧5) | | | | | | | |
|  | Men | | | | Women | | | |
| Rank | Age 20–29 | 30–39 | 40–49 | Over 50 | Age 20–29 | 30–39 | 40–49 | Over 50 |
| 1 | Stress reduction | Stress reduction | Better sleep | Better sleep | Better sleep | Better sleep | Better sleep | Better sleep |
| 2 | Better sleep | Better sleep | Stress reduction | Stress reduction | Stress reduction | Relaxation | Stress reduction | Relaxation |
| 3 | Job performance | Job performance | Work environment | Good lifestyle | Relaxation | Stress reduction | Stress coping | Stress reduction |
| 4 | Career | Good lifestyle | Job performance | Job performance | Good lifestyle | Problem solving | Relaxation | Work stress |
| 5 | Relaxation | Motivation | Positive feeling | Relaxation | Stress coping | Stress coping | Good lifestyle | Problem solving |
| 6 | Good lifestyle | Work environment | Relaxation | Work stress | Positive feeling | Work stress | Problem solving | Work environment |
| 7 | Motivation | Dairy behavior | Communication | Problem solving | Problem solving | Good lifestyle | Positive feeling | Positive feeling |
| 8 | Positive feeling | Positive feeling | Good lifestyle | Positive feeling | Job performance | Mindfulness | Job performance | Work-life balance |

*Note*. The personalized scenario was set based on the participants’ interests and preferences found in the preliminary survey among 1000 workers employed at micro-, small-, or medium-sized enterprises with less than 50 employees. Detailed information on how to personalize the scenario is described in Supplementary File 1.
